# Paediatric haematopoietic stem cell transplantation research priorities: results from an international multi-stakeholder Priority Setting Partnership

**DOI:** 10.64898/2026.03.18.26348770

**Authors:** Elizabeth Williams, Roxanne Dyas, Katherine Colman, Suzannah Kinsella, Amanda Gwee, Amy Lovell, Andrew R Gennery, Mary Slatter, Lori Chait-Rubinek, Eileen Van Der Stoep, Arjan Lankester, Hilda Mekelenkamp, Ben Gelbart, Karen Nicholson, Lorna McLeman, Shivanthan Shanthikumar, Vanessa Clifford, Theresa Cole, Gabrielle M Haeusler, Lisa M Ott de Bruin, Tim Prestidge, Adam Nelson, Kanchan Rao, Rachel Conyers, James Lind Alliance Priority Setting Partnership in Paediatric Haematopoetic Stem Cell Transplantation Steering Committee.

## Abstract

Co-designed research in paediatric HSCT is limited. We sought to determine research priorities which represent the shared priorities of patients, parents, carers, and healthcare professionals (HCP) within Australia, New Zealand, the Netherlands and United Kingdom.

An international, multiphase priority-setting methodology was implemented in partnership with the James Lind Alliance and delivered over an 18-month period. Part 1: an international scoping survey asked respondents to submit their research uncertainties related to paediatric HSCT. Part 2: summarising and evidence-checking the submitted uncertainties. Part 3: interim prioritisation survey. Part 4: consensus workshop.

In the first international scoping survey, 667 topic ideas were suggested (45% by consumers, 55% by HCP), which were categorized into 80 summary questions. After systematic literature review, 35 summary questions were judged to be true uncertainties (i.e. not answered by existing evidence). These 35 uncertainties were included in a second interim prioritisation survey, completed by 224 participants. From those, a shortlist of 19 questions was drawn. After a multistakeholder workshop, consensus was reached on the top 10 priorities.

The PSP identified important research gaps in the management of paediatric HSCT. Priority areas included: implementing personalised medicine approaches, improving immune recovery and adjunct interventions such as exercise, nutrition and microbiome-directed strategies.

## INTRODUCTION

Haematopoietic stem cell transplantation (HSCT) is a potentially curative therapy that replaces diseased or damaged bone marrow with healthy stem cells, restoring normal blood and immune system function. Since the first successful paediatric transplant in the late 1960s (1), HSCT has been used for a range of malignant and non-malignant conditions in children, including acute leukaemias, bone marrow failure syndromes, metabolic diosorders and immunodeficiencies (2). For many children, HSCT represents a final chance of survival, however it carries significant risk. Severe and varied complications, including infection, organ failure, and graft-versus-host disease (GvHD) remain common (3). Mortality rates in the first 3-6 months remain high (11% at 100 days, up to 30% at 6 months for patients requiring intensive care admission(4)). This early post-transplant period offers the clearest opportunity for research to reduce complications, enhance recovery, and improve survival.

Annually across Australia, New Zealand, the Netherlands, and the United Kingdom, approximately 750 children undergo HSCT (5–8). To date research agendas are predominantly shaped by clinicians, researchers, and government agencies, with minimal patient or family input (9). In paediatric HSCT key research consortia and collaborative groups publish research blueprints that strongly influence the field’s direction (10, 11). However, these frameworks and research priorities rarely include meaningful consumer perspectives, despite the growing attention to patient-reported outcomes and experiences (12, 13).

The James Lind Alliance (JLA) Priority Setting Partnership (PSP) model addresses this gap, uniting patients, carers, and health care professionals (HCPs) to identify shared research priorities (14). This is the first international application in paediatric HSCT (7, 15). The PSP developed across Australia, New Zealand the Netherlands and the United Kingdom establishes the top ten research priorities and provides an evidence-based direction to critical areas affecting survival, health outcomes and quality of life for paediatric HSCT recipients.

## METHODS

### Study design

This project adhered to the JLA methodology outlined in their guidebook (14). Ethics approval was granted by the Sydney Children’s Hospitals Network Human Research Ethics Committee (HREC) (2024/ETH01238). The PSP was conducted from January 2024 to June 2025 across Australia, New Zealand, the Netherlands and the United Kingdom due to similarities in healthcare systems and established collaborative networks. Research was led by The Murdoch Children’s Research Institute, Melbourne, Australia.

### Research Question

To identify and prioritise research questions relating to the first 180 days after paediatric HSCT

### Scope

This PSP focuses on priorities specific to paediatric HSCT within the first six months (180 days) following transplantation. All transplant indications were included (i.e. HSCT for malignant disease and genetically driven bone marrow failure disorders).

Eligibility for participation included (i) adolescents aged 13–<18 who had previously undergone an HSCT (either autologous or allogeneic) for any underlying condition, (ii) adults (>/=18 years) who had received an HSCT as a child, (iii) parents, carers, or family members of childhood HSCT recipients, (iv) HCPs involved in paediatric HSCT, and (v) individuals with professional or lived experience, or a demonstrated interest in the field and residing in Australia, New Zealand, the Netherlands, or the United Kingdom. The primary exclusion criteria were (i) living outside of those four countries, or (ii) being unable to complete the survey in English.

### Project management and process outline

The core management team, supported by a JLA Adviser, conducted the PSP in accordance with JLA methods and under the oversight of an international. Committee members were recruited through national HSCT networks and consumer advisory groups. The stepwise JLA process was then followed to identify the top ten priorities (Figure 1).

**Figure 1:**
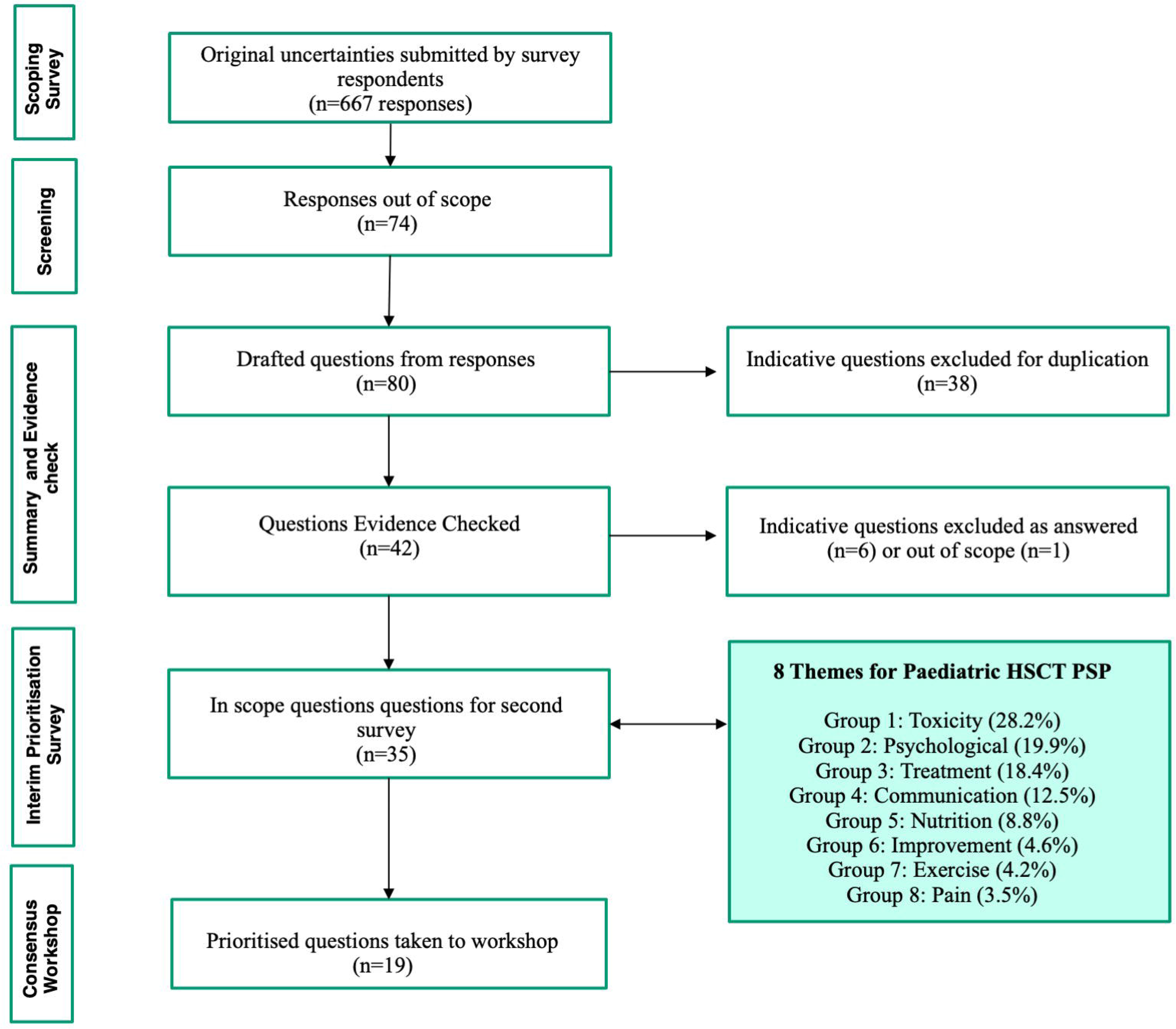
**Process Flow of the Paediatric Bone Marrow Transplant Priority Setting Partnership utilising the James Lind Alliance Methodology**

### PART 1: International Scoping Survey

The scoping survey captured stakeholder questions and ideas in a non-biased or leading manner by applying open-ended questions. Participants completed online acknowledgement of age-appropriate consent with an accompanying information statement prior to accessing the survey. Participation was anonymous with no identifying data collected. Demographic information was requested to monitor participation rates of target audiences (Table 1). Five healthcare professionals and five consumers across participating countries piloted the survey prior to public distribution and feedback was incorporated to improve user experience, including a trigger warning added to the survey introduction.

**Table 1:** Demographic data collected in scoping survey and Interim prioritisation survey. Abbreviations: HCP: Healthcare Professional; HSCT: Haematopoetic Stem Cell Transplant.

A promotional guide was developed by the lead researchers and shared with the Steering Committee and their networks. It outlined strategies to maximise survey participation, including public posters at clinical sites, online forums, social media engagement with advocacy groups, partnerships with philanthropic organisations, and presentations at academic and community events. Response rates and demographics were monitored to guide targeted promotion and support balanced participation.

### PART 2: Summarising and Evidence Checking

Responses from the scoping survey were assigned a unique identifier. Comments were reviewed and coded by two independent reviewers and grouped under corresponding themes (Supplementary 1). Coding discrepancies were resolved through collaborative discussion. Out of scope questions (defined as: questions specific to adult HSCT, questions not answerable by research, institutional specific, or long-term survivorship beyond 180 days post-transplant) were grouped separately and excluded from the priority setting process.

Following thematic analysis, overarching research questions were drafted from raw data responses to capture the core uncertainties expressed, while remaining true to the original wording and intent of contributors (Supplementary 2). After review by subgroups of the Steering Committee, consolidated questions written in language accessible to non-medical audiences were taken to the evidence checking phase.

A literature review was conducted to determine whether each summary question represented a true evidence uncertainty. Researchers (termed ‘information specialists’ according to JLA methodology) conducted the evidence check accordingly with the question verification form (Supplementary 3) and following predefined criterion (Table 2).

**Table 2:** Criteria for evaluating existing evidence for research questions. If all criteria were met (i-iv) the evidence was considered answered. If no criteria were met the question was unanswered. If some of the criteria were met the question was considered partially answered.

To classify a completely answered research question the following criteria were defined:

(i) Original research published in high-quality publication (systematic review, clinical guideline, or meta-analysis); AND
(ii) publication within the last five years (2020–2025); AND
(iii) relevance to paediatric HSCT.

If one or two criteria were met, the question was classified as *partially answered*, and if none were met, the question was classified as *unanswered*. All unanswered and partially answered questions progressed to the second survey.

### PART 3: Interim Prioritisation Survey

Survey two aimed to prioritise the research uncertainties identified in the scoping survey. Online consent was required, and all responses were anonymous with no identifying data collected. Participation in survey one was not a prerequisite. Questions were presented in a randomised sequence using REDCap® to minimise order bias, and participants selected up to ten questions they viewed as most important for future research. The interim prioritisation survey was promoted using the same multimodal strategies as survey one, with real-time demographic monitoring used to tailor promotion and support balanced participation.

Responses were analysed by country (Australia, New Zealand, the Netherlands and the United Kingdom) and stakeholder group (family member, recipient, HCP). A tally system generated priority scores and identified the top ten questions for each group. These combined scores formed the interim workshop shortlist of 19 questions, which the Steering Committee reviewed to ensure representation of all stakeholder priorities including minority groups.

### PART 4: Cosensus Workshop

Four trained JLA Advisers facilitated the workshop which employed the JLA’s adapted Nominal Group Technique to ensure balanced, inclusive discussion (14). Over two half-day sessions, participants worked in four diverse, facilitated groups to deliberate and rank the 19 questions. Combined rankings produced the final list and top ten priorities.

## RESULTS

### Project design and management

Oversight for the project was provided by a Steering Committee of 31 members. The committee represented four countries: Australia (14), New Zealand (two), the Netherlands (four), and United Kingdom (11), and comprised 23 HCPs, seven family members of children who had received an HSCT, and one individuals who had undergone an HSCT as a child. Among HCPs, 65% were medical doctors (seven sub-specialties) 35% were allied health professionals (4 subspecialities).

### PART 1: International Scoping Survey

This survey, completed by 148 people between 8 August and 31 October 2024, generated 667 unique questions or comments related to childhood HSCT. Balanced input distribution was achieved, with 55% of respondents being HCPs and 45% consumers (Figure 2A). The remaining respondents included one charity representative and three who did not disclose their background. Geographically, respondents were from Australia (38.5%), United Kingdom (32.4%), Netherlands (22.3%) and New Zealand (6.8%) (Figure 2B). Reported ancestry distribution revealed European (67.1%), Oceanian (16.4%) and East Asian (6.8%). Various other ancestral groups contributed less than 3% each (Figure 2C).

**Figure 2:**
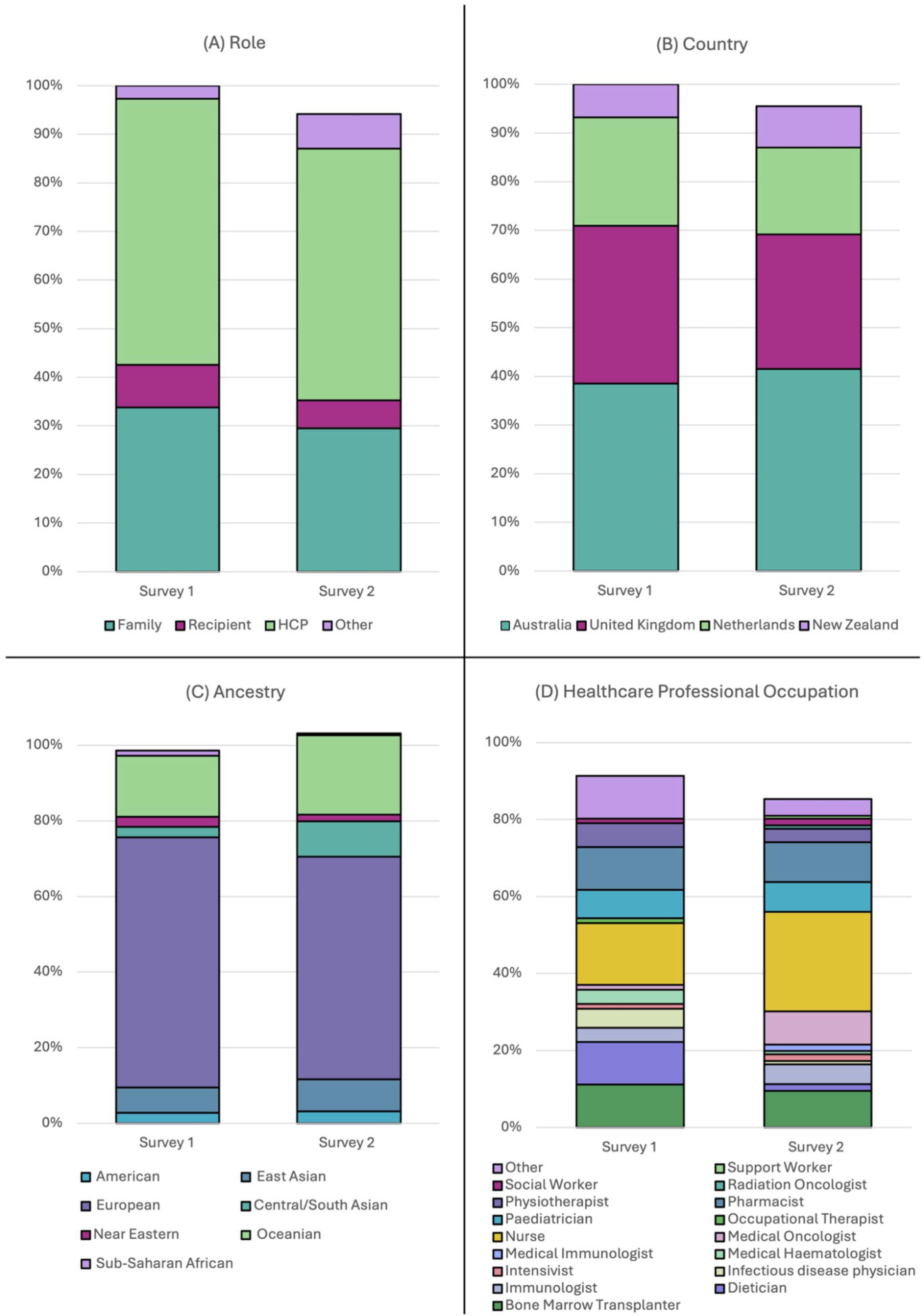
Demographics distribution of International Scoping Survey (Survey 1) and Interim Prioritisation Survey (Survey 2) respondents by role (A), country (B), ancestry (C), and healthcare professional occupation (D).

### PART 2: Summarising and Evidence Checking

Initial review prompted the removal of 74 comments deemed out of scope. Thematic analysis categorised the remaining 593 data points into eight key themes: (i) toxicity and side effects 28.2%, (ii) psychological wellbeing and support 19.9%, (iii) standardisation of treatment 18.4%, (iv) communication 12.5%, (v) nutrition 8.8%, (vi) exercise and rehabilitation 4.2%, (vii) improvement of processes 4.6%, and (viii) pain management 3.5% (Figure 3A).

**Figure 3:**
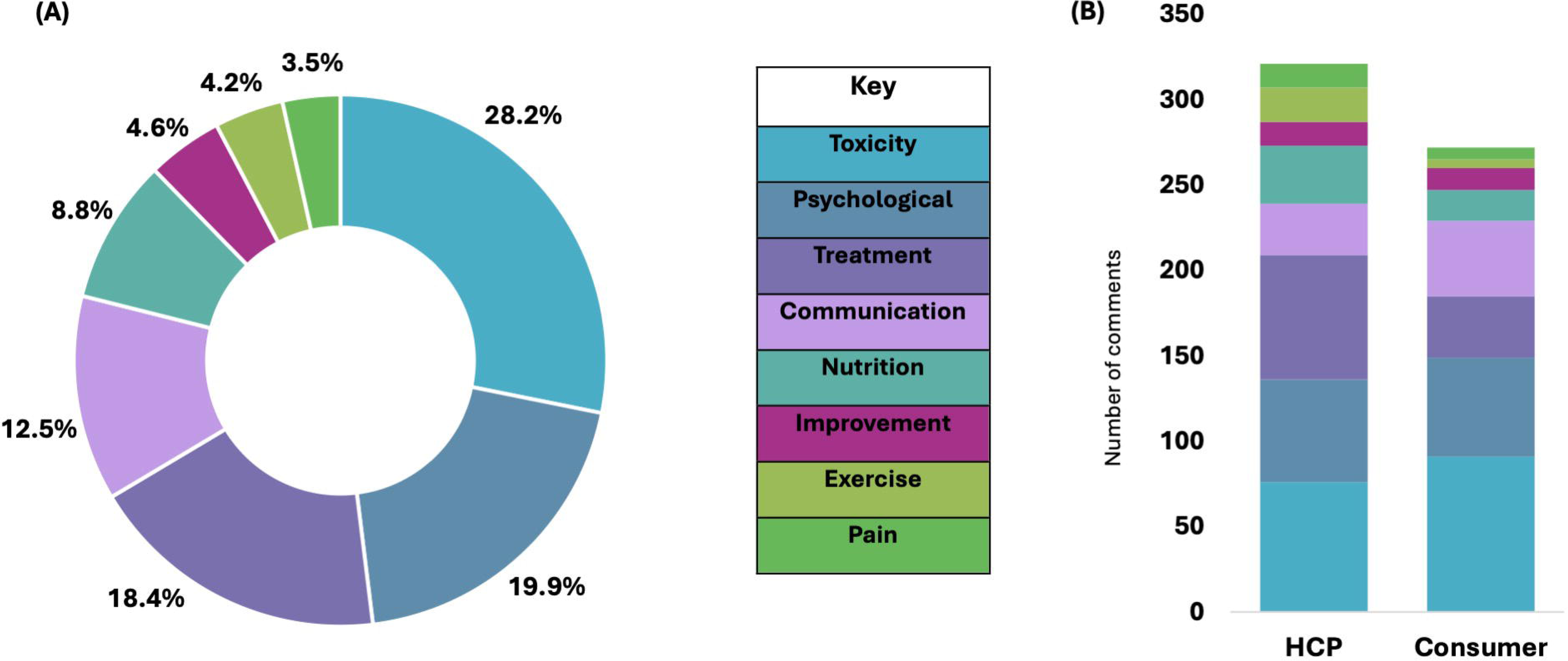
Distribution of thematic analysis of responses. (A) Proportion of all submitted questions and comments mapped to each theme. (B) Number of responses attributable to each theme for healthcare professionals versus consumers. Abbreviations: HCP: Healthcare professional.

Comparing the 321 comments from healthcare professionals and 272 comments from consumers, differences in topic prioritisation were observed (Figure 3B). Healthcare professionals more frequently prioritised standardisation of treatment (22.7 versus 13.2)*, nutrition (10.6 versus 6.6)*, exercise and rehabilitation (6.2 versus 1.8)*, and pain (2.4 versus 1.6)*. Consumers were more likely to prioritise toxicity and side effects (33.5 versus 23.7)*, psychological wellbeing and support (21.3 versus 18.7)*, communication (16.2 versus 9.4)* and improvement of processes (4.8 versus 4.4)* (*Percentages after normalisation).

Following the JLA question verification process of 42 questions, 35 questions were identified as unanswered or partially answered by existing research and progressed to survey two for prioritisation.

### PART 3: Interim Prioritisation Survey

A total of 224 respondents completed this survey between 2 April and 2 June 2025. Input was balanced between healthcare professionals (55%) and consumers (45%). Geographically, respondents were primarily from Australia (41.5%), the United Kingdom (27.1%), the Netherlands (22.4%), and New Zealand (7.6%), with 1.4% not specifying their location. Reported ancestry distribution included 58.9% European, 20.3% Oceanian, 9.1% Central/South Asian, 8.2% East Asian, 3% American with other ancestral groups contributing less than 3% (Figure 2C). While patients and clinicians showed some alignment in their rankings, differences in emphasis were nevertheless evident, highlighting the importance of understanding priorities across a range of stakeholders. Clinical intervention topics such as diagnosing and treating thrombotic microangiopathy (TMA) were prioritised more highly by HCPs than by recipients and family members (ranked 7 versus 28 and 33, respectively). In contrast, quality-of-life issues, such as maintaining schooling and education during transplantation, were ranked higher by recipients and families (2 and 4) than by HCPs (32) (Supplementary 4). The results informed the selection of questions taken forward to the final priority-setting workshop.

### PART 4: Consensus Workshop

Thirteen HCPs and nine consumers, from Australia (ten), New Zealand (four), the Netherlands (three), and the United Kingdom (five) participated in the workshop. Eight observers from philanthropic organisations and research centres attended but did not contribute. Three additional consumers were unable to participate on the day. Four JLA Advisers facilitated discussions, employing the Nominal Group Technique to reach consensus on the top ten research priorities for paediatric HSCT (Supplementary 5).

The final top 10 research questions are summarised in Table 3, with detailed rankings across stakeholder groups available in Supplementary 6.

**Table 3:** Top Ten Research Priorities for Paediatric HSCT. Abbreviations: GvHD: Graft Versus Host Disease; HSCT: Haematopoetic Stem Cell Transplantation.

## DISCUSSION

This PSP is the first international initiative to systematically define paediatric HSCT research priorities representing a shared vision between HCP and consumers. The top 10 priorities integrate therapeutic optimisation with proactive supportive-care strategies across the biological, functional and psychosocial dimensions of paediatric HSCT.

The paediatric HSCT research landscape remains largely shaped by clinician and researcher-driven agendas (7, 15), with limited integration of consumer perspectives. At the same time, the overall volume of paediatric HSCT clinical-trial driven research is low; with only eight investigator-initiated or commercial-led interventional trials registered on clinicaltrials.gov between 2020 and 2025. Moreover, as this PSP demonstrates, consumers and HCPs often have differing perspectives and priorities, highligting the imperative of seeking input across a wide range of stakeholders. This combination of challenges underscores the need for coordinated prioritisation and reinforce the value of this PSP in establishing a rigorous, patient-centered research agenda.

Treatment-related risk remains a major determinant of paediatric HSCT outcomes. Registry data from the Center for International Blood and Marrow Transplant Research and European Society for Blood and Marrow Transplantation consistently show that toxicities, including GvHD, infection, and organ injury, continue to drive morbidity and mortality despite improvements in supportive care (3, 8, 16). Reflecting this burden, toxicity-related questions comprised the largest proportion of submissions in this PSP and ranked prominently in the final prioritised list. Stakeholders emphasised the need to better understand, predict and prevent treatment-related toxicities, highlighting both the limitations of current approaches and the emerging potential of personalised strategies such as pharmacogenomics (17, 18), immune profiling (19) and pharmacokinetic-guided dosing (20). In paediatric HSCT, early precision-medicine efforts are already evident, for example, model-informed dosing of busulfan in children utilises weight, age and pharmacokinetics to optimise exposure and reduce toxicity (21). Parallel work on treosulfan has demonstrated significant inter-patient variability in pharmacokinetics and suggests the need for targeted first-dose strategies in paediatric HSCT (22). Finally, serotherapy agents such as alemtuzumab are also entering personalised dosing, with exposure–response modelling in children beginning to link drug levels to immune recovery and transplant outcomes (23, 24). Combined with expanding pharmacogenomic knowledge (25), these approaches illustrate a shift toward evidence-driven, individualised conditioning and immune-modulation strategies, with recognition of this priority shared by consumers and clinicians to reduce treatment toxicity.

Beyond treatment-related toxicity, several priorities reflected the central biological challenges of graft function and immune reconstitution. Delayed or incomplete immune recovery is a well-established driver of post-transplant morbidity and mortality (16, 26), increasing susceptibility to opportunistic infections (27) and prolonging psychosocial and physical restrictions. Within this context, GvHD remains a persistent determinant of both early toxicity and long-term recovery, with registry data identifying it as a major contributor to transplant-related mortality and chronic morbidity (6, 8, 16). Stakeholders emphasised the need for improved prediction, prevention and early identification of GvHD, including the potential roles of serotherapy optimisation, biomarker-guided risk stratification and enhanced immune profiling. Consistent with these biological challenges, stakeholders prioritised questions addressing immune restoration, graft failure, infection prevention strategies and the optimisation of isolation practices (Priorities 4, 5, 7, 8). The importance of gut microbial diversity also emerged, aligning with growing evidence linking microbiome disruption to immune dysregulation, infection risk and GvHD (9). Although microbiome-directed therapies such as faecal microbiota transplantation remain experimental in children, these priorities highlight strong interest in mechanistic pathways that may accelerate immune recovery, reduce alloreactivity and stabilise graft function. Together, these themes underscore the need for integrated, biologically informed strategies to improve early post-transplant recovery.

Alongside biological determinants of outcome, stakeholders highlighted the importance of supportive-care and functional domains that influence recovery trajectories in paediatric HSCT. Sarcopenia (loss of muscle mass), impaired physical function and nutritional deficits are increasingly recognised as modifiable risk factors associated with delayed engraftment, higher infection rates, prolonged hospitalisation and poorer survival (9, 13). Emerging data from paediatric oncology and small HSCT cohorts indicate that structured exercise programs and nutritional optimisation are feasible and may mitigate early deconditioning and longer-term frailty, which affects an estimated 7% of paediatric transplant survivors (28–30). Despite this, prehabilitation strategies remain underutilised. The inclusion of exercise and nutritional optimisation within the top priorities reflects the growing recognition amongst clinicians and families of improving physical resilience before and after HSCT (28, 30).

Psychosocial and neurocognitive outcomes also emerged as major priorities, reflecting the substantial emotional and developmental burden associated with paediatric HSCT. Psychological distress affects one third of HSCT recipients and is associated with a poorer quality of life, impaired treatment adherence and delayed recovery (31). Families similarly experience high levels of anxiety, uncertainty and caregiver strain thorughout the transplant trajectory. Cognitive impairments including deficits in attention, processing speed and memory, are well documented late effects and may limit educational and social reintegration (32). Stakeholders therefore emphasised the need for improved identification of children at risk of psychological and cognitive difficulties, along with evidence-based interventions to support coping, resilience and neurocognitive development.

A major strength of this PSP is its multinational scope, diverse Steering Committee (Supplementary 7), and rigorous JLA methodology, ensuring transparent and balanced engagement between HCP, patients and caregivers. The resultant set of priorities are both globally relevant and sufficiently granular to inform local clinical practice and research planning. Importantly, the PSP actively engaged established stakeholders across the paediatric HSCT and paediatric oncology ecosystem (e.g. Anthony Nolan Trust and Bubble Foundation (United Kingdom), Maddie Riewoldt’s Vision and the Australian and New Zealand Children’s Haematology Oncology Group (Australia)) enhancing the legitimacy and reach of the final priorities. Through these partnerships, this PSP can be utilised by funding agencies, trial consortia and advocacy organisations to shape future investment.

This PSP has several limitations that should be considered when interpreting the findings. First, although multinational, participation was greatest in Australia and the United Kingdom, reflecting established collaborator networks and potentially introducing regional weighting in priority selection. Broader representation from low and middle-income settings, where indications, access to transplant and supportive care differ substantially, may have yielded additional perspectives. Second, this PSP focussed only on the first 180 days after HCT and may exclude late complications. Third, surveys were written in English, which may have limited engagement from non-English-speaking families and clinicians. Fourth, while the JLA methodology ensures transparent and equitable stakeholder input, its reliance on consensus means that highly specialised or emerging research areas may not reach the final prioritised list despite their scientific importance. Finally, as with all PSPs, prioritisation reflects the views of those who participated; certain professional groups, including late-effects specialists, psychologists and allied health professionals, were under-represented relative to transplant physicians and nurses. Increased participation from transplant recipients and Indigenous consumers may also have broadened applicability of the identified priorities.

This PSP provides the first internationally derived, stakeholder-informed research agenda for paediatric HSCT. The priorities identified span biological, functional and psychosocial domains, reflecting the complexity of transplant care and the need for coordinated, multidisciplinary approaches to improve outcomes. By consolidating the perspectives of clinicians and consumers, the PSP offers a clear framework to guide research investment, catalyse trial development and support the translation of emerging personalised and supportive-care strategies into clinical practice.

## Data Availability

Data is publically available via the James Lind Alliance Priority Setting Partnership Website.

https://www.jla.nihr.ac.uk/priority-setting-partnerships/paediatric-bone-marrow-transplant#tab-key-documents

## ACKNOWLEDGEMENTS

We acknowledge the contributions of the James Lind Alliance, Partner organisations (Supplementary 6) and Steering Committee (Supplementary 7).

## AUTHOR CONTRIBUTIONS

RC conceived, and with EW and RD designed the study compliant with JLA methodology. EW and RD collected and analysed the data. EW and RC drafted the manuscript. AG, MS, KC, EV, AL, SS, AG, LCR,KN, BG, MC, SK, LD, AL, HM, LM, GH, TC, AN, TP, KR reviewed, edited and approved the final manuscript.

## DATA AVAILABILITY STATEMENT

The datasets are available from the corresponding author on request.

## ETHICS APPROVAL STATEMENT

This project was approved by The Royal Children’s Hospital, Human Research Ethics Committee HREC/105834.

