## Supplemental Files for "Paediatric haematopoietic stem cell transplantation research priorities: results from an international multi-stakeholder Priority Setting Partnership"

Supplementary Files

**Index**

| Supplementary Files | Topic | Page |
| --- | --- | --- |
| Table S1 | Thematic Allocation Guide | 2 |
| Table S2 | Summary Question Generation | 3 |
| Table S3 | Question Verification Form | 4 |
| Table S4 | Survey 2 research question ranking scores by stakeholder and country groups | 5 |
| Table S5 | Workshop question rankings using the Nominal Group Technique | 6 |
| Table S6 | Partnering organisations | 7 |
| Table S7 | Paediatric HSCT Steering Committee Members | 8 |

| Guide for Allocating Comments and Questions into Themes | | |
| --- | --- | --- |
| Theme / Category | **Theme Focus** | **Key areas** |
| Standardisation and Treatment | Treatment options and selections for HSCT recipients (excluding side effects) | Guideline  Immune reconstitution  Treatment  Prophylaxis  Risk assessment  Donor risks and requirements  Conditioning options |
| Toxicity & side-effects | Managing side effects and resulting complications | Management of treatment related symptoms and side effects including:  Mucositis  GVHD  TMA  Infection  Complications of treatment  Fertility |
| Communication | Improving how information is communicated and shared among healthcare providers, patients, and families in childhood cancer care. | Clinician-patient/family communication  Multidisciplinary team communication  Access to understandable information  Communication in decision-making  Management of expectations  Consent |
| Psychological wellbeing and support | Emotional, mental, and social needs of children requiring a HSCT and their families | Mental health support and services for families  Mental health support and services for families - post discharge  Mental health support and services for families - hesitant  Mental health support and services for children/patients  Isolation  Peer Support  Sexuality/Puberty  Schooling  Cultural |
| Nutrition | Ways to ensure the best nutrition for children requiring a BMT  Optimising nutritional status and microbiome  Best options for children having a BMT | Gut microbiome  Best forms of nutrition support  Nutrition affecting BMT outcomes  TPN v enteral feeds  Different types of feeds  Assessment of nutrition status  Feeding guidelines |
| Exercise and Prehabilitation/ Rehabilitation | Ways to implement and promote physical activity for BMT recipients | Structured exercise programs  Access to equipment  Implementation of activity in day-to-day inpatient stay |
| Pain | Pain management | Treatment options and management  How pain affects sleep/mental health/physical function |
| Improvements | Ways to improve care experience and training while in hospital | Training  Discharge planning  Holistic care wishes |

**Supplementary 1: Question and comment allocation guide.** Abbreviations: BMT Bone Marrow Transplantation; TA-TMA: Transplant Associated Microangiopathy; TPN: Total Parenteral Nutrition.

| Example of Survey Questions Categorised and Synthesised into Summary Questions | | | |
| --- | --- | --- | --- |
| THEME:  Toxicity and Side Effects | | | **SUMMARY QUESTION TS7:**  **How can mucositis be prevented and treated in transplant?** |
| FOCUS:  Mucositis | | |  |
| RECORD | **WHO** | **Q. ID** | **RESPONSE:** |
| 12 | HCP | 6.4 | novel management mucositis |
| 33 | FAMILY | 8.5 | I wish that I had asked more questions about the mucositis that was expected about anything that we could do to help |
| 33 | FAMILY | 8.6 | Or that I had asked about other treatments for this *(mucositis)* such as helios (that seems very common in the USA) |
| 133 | HCP | 47.3 | I have questions about best feed during mucositis. Can different type of feed make a difference? |
| 165 | HCP | 61.3 | How do we optimize mucositis care based on best practices and evidence. |
| 165 | HCP | 61.7 | How do we optimize mucositis care, interventions for nauseousness, tube / parenteral feeding, graft versus host treatment and create a more uniform approach, based on best practices and evidence. |
| 328 | REC | 110.3 | I wasn't prepared for the mucositis and not being able to eat because of it |
| 358 | HCP | 123.4 | Nausea and vomiting management in the context of mucositis |
| 378 | FAMILY | 130.1 | Side effects and complications. Why isn't there something to help with Mucositis? |
| 393 | FAMILY | 135.2 | I wish I had known that the morphine and ketamine would mean my son doesn't remember how agonisingly painful the mucositis was. |
| 174 | HCP | 65.2 | The impact of mucositis and gut GVHD on bacterial and fungal infection risk is profound. More collaborative research is need to identify strategies to limit the impact of this. |
| STEERING COMMITTEE ACTION/SUGGESTION: How can mucositis be better managed in transplant? | | | |
| Legend  HCP: Healthcare professionals  Family: Recipient of a haematopoietic stem cell transplantation as a child, family members or carers  GVHD: Graft versus host disease  TS7: Question code (Toxicity and Side effects question 7) | | | |

**Supplementary 2:** Example survey question categorisation approach to form summary questions. Abbreviations: GVHD: Graft Versus Host Disease; HCP: Healthcare Professional; TS7: Question code (Toxicity and Side Effects Question 7).

**Question Verification Form**

**Name of the PSP**

Bone Marrow Transplantation in Paediatrics Priority Setting Partnership

**Scope of the PSP**

The aim of the Bone Marrow Transplantation (BMT) in Paediatrics PSP is to identify the unanswered questions about BMT and its resultant complications from conditioning until Day 180 post BMT from the shared perspectives of adolescents and young adults who have received BMT, their caregivers, physicians and support personnel, researchers, patient engagement specialists and/or members of patient organisations, and then prioritise those unanswered questions that these groups agree are the most important for research to address.

**Overview of approach to checking whether the questions were unanswered**

In partnership with the Steering Group, all questions and comments submitted from respondents from the first PSP survey were collated into themes and then summary questions.

To evaluate if these summary questions are unanswered by previous research, a literature search of each finalised summary question (that is in-scope) will be undertaken and be categorised as being:

1. Answered: Reliable, up-to-date systematic reviews, meta-analyses or clinical guidelines has already been published.
2. Partially answered:

- Relevant, reliable and up-to-date systematic reviews, meta-analyses and evidence-based guidelines but do not address continuing questions.
- Relevant systematic reviews but not up to date (published before 2020).
- Current clinical trial but not exhaustive enough to answer the question.

3. Not answered: No relevant systematic review or clinical guideline identified.

All questions categorised as ‘2’ or ‘3’ will be deemed unanswered and taken to interim priority setting.

**The type(s) of evidence used to verify your questions as unanswered**

We will verify questions as unanswered when considering the following evidence types:

- Systematic reviews (which had reviewed randomised controlled trials (RCTs), quasi RCTs, cluster RCTs, cohort studies, cross-sectional studies, qualitative literature)
- Evidence-based national guidelines
- Clinical trials: clinical trials will be used to identify questions that may be answered in the coming years. An ongoing study may not mean that an uncertainty will be addressed. Steering group members will discuss ongoing studies and document any decisions made because of information found.

**Sources searched to identify that evidence**

The following searches will be undertaken as relevant for each question:

- The Cochrane Library
- Epistemonikos
- Databases:  Medical Literature Analysis and Retrieval System Online (MEDLINE), Cumulative Index to Nursing and Allied Health Literature (CINAHL), PubMed (U.S. National Library of Medicine), Turning Research Into Practice (TRIP) Database, BMJ Best Practice, ClinicalKey (Elsevier),
- European guidelines: National Institute for Health and Care Excellence (NICE), the European Society for Medical Oncology (ESMO) guidelines
- Clinical Trials Registry: The World Health Organization International Clinical Trials Registry Platform (ICTRP), the Australian New Zealand Clinical Trials Registry (ANZCTR), and ClinicalTrials.gov (U.S. National Library of Medicine).

**Search terms used?**

For our search on Cochrane Reviewers and database searches, we will include:

systematic review* OR meta-analysis OR meta analysis* OR guideline* OR review*

**AND**

child OR adolescent OR pediatric* OR paediatric* OR teen* OR young* OR childhood

**AND**

Stem cell transplant OR bone marrow transplant OR hemopoietic stem cell transplant OR haemopoietic stem cell transplant OR cord blood transplant OR autologous OR allogeneic

**AND**

*additional search terms to reflect the concepts/focus of each summary question*

**Parameters of the search (eg time limits, excluded sources, country/language) and the rationale for any limitations**

Time limits

Searches will be limited to evidence published from January 2020 to January 2025. Where we are aware that older evidence has answered a question, and further research or an update may be unnecessary, this evidence will be presented to the Steering Group, with a consensus reached on whether the question is considered ‘answered’ or ‘partially answered’.

Country and language

- Searches will be limited to publication in English in a peer-reviewed journal (or peak body, in the case of guidelines).
- Evidence from any country that has comparable health care systems will be accepted.

Sample

Scenarios where we find evidence that includes young adults (in the age group of ≥19 and <25 years), along with adolescents that are expected to be available in literature those studies will be considered.

Similarly, there may be scenarios where broader literature in other genetic conditions may be included. These scenarios will be discussed with the Steering Group.

In addition to the literature review, the Steering Group will be asked to confirm the availability of any additional evidence within their expertise as necessary.

**Supplementary 3: Evidence Checking Verification Form.** Abbreviations: BMT: Bone Marrow Transplantation; PSP: Priority Setting Partnership

|  | **Ranking Scores** | | | | | | | |
| --- | --- | --- | --- | --- | --- | --- | --- | --- |
| **Question** | **Average** | **Family** | **Recipient** | **HCP+ ORG** | **ND** | **AUS** | **UK** | **NZ** |
| What are the psychological concerns and needs of patients and families at different times during transplant? | 2.67 | 2 | 1 | 5 | 1 | 5 | 2 | 4 |
| What are the causes of early relapse after transplant, and how can this information help identify high-risk patients and develop strategies to prevent relapse? | 3.67 | 1 | 6 | 4 | 15 | 2 | 1 | 4 |
| How can immune function be restored more rapidly following transplantation? | 5.33 | 9 | 5 | 2 | 15 | 1 | 11 | 2 |
| How can treatment related toxicities best be predicted and reduced in transplant patients? | 6.00 | 6 | 11 | 1 | 7 | 3 | 2 | 26 |
| How can a personalised approach to medication reduce adverse effects and improve recovery and outcomes for transplant patients? | 7.33 | 9 | 6 | 7 | 3 | 12 | 8 | 26 |
| Sometimes, transplanted bone marrow doesn't work properly. How do you improve its function or manage graft failure? | 7.33 | 4 | 11 | 7 | 7 | 4 | 2 | 32 |
| What are the most effective ways to prevent, identify and treat GVHD? | 9.00 | 6 | 18 | 3 | 5 | 6 | 7 | 1 |
| How does a transplant impact cognition, memory and the way your think, and can this be improved or prevented? | 9.00 | 3 | 2 | 22 | 10 | 8 | 6 | 19 |
| What are the best exercises to maintain muscle and manage fatigue. How can these be used by transplant patients? | 10.00 | 13 | 2 | 15 | 9 | 20 | 16 | 4 |
| How can we maximise the effectiveness of medicines and tests in preventing, detecting and treating infections in transplant patients? | 11.33 | 12 | 6 | 16 | 21 | 8 | 10 | 11 |
| What are the best ways (pharmaceutical and non-pharmaceutical) to control and manage pain associated with transplant? | 11.33 | 16 | 11 | 7 | 3 | 15 | 20 | 19 |
| How can schooling and education be best maintained for children undergoing transplant? | 12.67 | 4 | 2 | 32 | 15 | 20 | 5 | 26 |
| How can genetic testing predict the transplant patients most at risk of severe toxicity? | 13.33 | 15 | 18 | 7 | 15 | 8 | 14 | 13 |
| How can including peer-to-peer support networks throughout transplant, reduce social isolation and improve emotional outcomes? | 13.67 | 8 | 11 | 22 | 5 | 19 | 12 | 26 |
| What are the necessary infection prevention and isolation measures? | 14.00 | 16 | 6 | 20 | 21 | 20 | 16 | 26 |
| How can vital organ failure be identified, prevented, and managed in transplant recipients? | 15.00 | 9 | 6 | 30 | 21 | 15 | 8 | 34 |
| How can mucositis be better managed in transplant? | 15.33 | 16 | 18 | 12 | 21 | 13 | 20 | 2 |
| Research shows that multidisciplinary care improves the well-being of patients and their families. How can this be implemented in transplant? | 15.33 | 16 | 18 | 12 | 2 | 24 | 20 | 19 |
| What is the role of a diverse intestinal microbiome in transplant outcomes, and how can this diversity be achieved? | 17.67 | 29 | 18 | 6 | 15 | 6 | 26 | 19 |
| How can VOD be predicted and prevented? | 18.33 | 22 | 11 | 22 | 27 | 24 | 30 | 4 |
| Can discharge planning be improved through enhanced inter-hospital communication and family training? | 20.67 | 20 | 11 | 31 | 27 | 30 | 16 | 13 |
| What are the best ways to address the cultural needs of children and families in transplant care? | 20.67 | 26 | 18 | 18 | 12 | 33 | 16 | 13 |
| Can standardised conditioning approaches be improved and personalised? | 20.67 | 14 | 29 | 19 | 21 | 18 | 12 | 19 |
| When should CMV prophylaxis be used, and what is the medicine of choice? | 21.33 | 26 | 18 | 20 | 31 | 20 | 26 | 11 |
| Can graft manipulation and cellular therapy be used to reduce the risk of viral infections in transplant patients? | 21.67 | 33 | 18 | 14 | 33 | 14 | 20 | 19 |
| What role does nutritional support and diet play in GVHD? | 22.33 | 22 | 18 | 27 | 15 | 26 | 32 | 4 |
| What is the best way to diagnose and treat TMA? | 22.67 | 33 | 28 | 7 | 33 | 8 | 28 | 13 |
| How does implementing multidisciplinary rehabilitation influence clinical outcomes and support reintegration into the community? | 22.67 | 22 | 18 | 28 | 12 | 26 | 28 | 34 |
| What are the most effective methods for assessing and providing nutrition and energy needs in children during transplant? | 23.33 | 15 | 29 | 26 | 14 | 30 | 14 | 4 |
| What causes taste disturbances in transplant patients, and can anything help with this? | 23.67 | 26 | 11 | 34 | 30 | 35 | 32 | 4 |
| How can antifungal prophylaxis and treatment be improved to lower the risk of infection? | 24.33 | 20 | 28 | 25 | 26 | 26 | 20 | 13 |
| How can informed consent and involvement in decision making about the transplant process be improved for children and families? | 24.33 | 29 | 28 | 16 | 10 | 15 | 30 | 19 |
| Are transplant treatment guidelines followed, and how does this impact patient outcomes? | 26.67 | 22 | 29 | 29 | 27 | 26 | 25 | 32 |
| Is there always an association between graft-versus-leukemia effect and graft-versus-host disease in transplant? | 31.00 | 31 | 29 | 33 | 31 | 30 | 34 | 13 |
| How can we improve the treatment of extramedullary leukaemia in transplant patients? | 31.67 | 32 | 28 | 35 | 35 | 34 | 35 | 26 |

**Supplementary 4 : Ranking of Survey 2 questions subdivided according to country or respondent.** Abbreviations: AUS: Australia; CMV: Cytomegalovirus; GVHD: Graft Versus Host Disease; NDL: Netherlands;NZ: New Zealand; TMA: Thrombotic Microangiopathy; UK: United Kingdom.

| **Workshop question ranking using the Nominal Group Technique** | | | | | |
| --- | --- | --- | --- | --- | --- |
| **Ranking in groups** | | | | **Question** | **Overall Rank** |
| **G1** | **G2** | **G3** | **G4** |  |  |
| 1 | 1 | 1 | 1 | How can a personalised approach to treatment and medication (e.g. genetic testing) reduce adverse effects and improve recovery and outcomes for transplant patients? | 1 |
| 2 | 2 | 2 | 5 | How can treatment related toxicities best be predicted and reduced in transplant patients? | 2 |
| 3 | 3 | 3 | 4 | What are the most effective ways to prevent, identify and treat GvHD? | 3 |
| 4 | 4 | 4 | 2 | How can immune function be restored more rapidly following transplantation? | 4 |
| 6 | 5 | 5 | 3 | What is the role of a diverse intestinal microbiome in transplant outcomes, and how can this diversity be achieved? | 5 |
| 12 | 6 | 6 | 6 | What are the psychological concerns and needs of patients and families at different times during transplant? | 6 |
| 8 | 8 | 10 | 9 | Sometimes, transplanted bone marrow doesn't work properly. How do you improve its function or manage graft failure? | 7 |
| 5 | 7 | 11 | 13 | What are the necessary infection prevention and isolation measures? | 8 |
| 9 | 9 | 9 | 10 | What are the best exercises to maintain muscle and manage fatigue. How can these be used by transplant patients? | 9 |
| 13 | 13 | 7 | 8 | How does a transplant impact cognition, memory and the way your think, and can this be improved or prevented? | 10 |
| 11 | 11 | 8 | 14 | What are the best ways (pharmaceutical and non-pharmaceutical) to control and manage pain associated with transplant? | 11 |
| 7 | 10 | 13 | 15 | How can we maximise the effectiveness of medicines and tests in preventing, detecting and treating infections in transplant patients? | 12 |
| 14 | 14 | 14 | 7 | How can vital organ failure be identified, prevented, and managed in transplant recipients? | 13 |
| 10 | 12 | 15 | 12 | What are the causes of early relapse after transplant, and how can this information help identify high-risk patients and develop strategies to prevent relapse? | 14 |
| 16 | 15 | 16 | 11 | Research shows that multidisciplinary care improves the well-being of patients and their families. How can this be implemented in transplant? | 15 |
| 17 | 17 | 12 | 17 | What are the best ways to address the cultural needs of children and families in transplant care? | 16 |
| 15 | 16 | 17 | 16 | How can including peer-to-peer support networks throughout transplant, reduce social isolation and improve emotional outcomes? | 17 |
| 18 | 18 | 18 | 18 | What is the best way to diagnose and treat TMA? | 18 |
| 19 | 19 | 19 | 19 | How can schooling and education be best maintained for children undergoing transplant? | 19 |

**Supplementary 5: Question rankings from each of the four workshop groups.** The rank from each group was combined by majority to achieve the overall rank. Position were classified as gold (questions 1-10); silver (11-15); bronze (16-19) to assist in visual representation during workshop grading. Abbreviations: GVHD: Graft Versus Host Disease; TMA: Thrombotic Microangiopathy.

**Partnering organisations**

Australian and New Zealand Children’s Oncology Group (ANZCHOG) and sub-group TACTIC (national malignant discussion BMT group)

Zero Childhood Cancer Program Patient Advisory Group (ZERO PAG)

Australian Society of Clinical Immunology and Allergy (ASCIA) and the sub-group TAPID (national immunology BMT discussion group)

Australia and New Zealand Transplant & Cellular Therapies (The Blood and Marrow Transplant Advisory Committee)

Immunodeficiency UK

Anthony Nolan | Saving lives through stem cells

British Society of Blood and Marrow Transplantation and Cellular Therapy

Youth Cancer Advisory Board and Youth voice

Childhood Cancer Association

Arrow: The Bone Marrow Transplant Foundation

Bubble Foundation

Children's Cancers Foundation

The Kids Cancer Project

Leukaemia & Blood Cancer New Zealand

Maddie Riewoldt's Vision

Paediatric Study Group - ANZICS

Young Lives Vs Cancer

Children's Cancer CoLab.

**Supplementary 6: Partner organisations.** The partner organisations agreed and participated in the priority setting partnership by disseminating Survey 1 and Survey 2.

| **Steering Committee Members** | | |
| --- | --- | --- |
| **Member Name** | **Country** | **Role** |
| Abbie YOUNG | The United Kingdom | Consumer Representative (Adolescent and Young Adult HSCT Recipient) |
| Adam NELSON | Australia | Paediatric Oncologist and Transplant and Cellular Therapy Physician |
| Aleksandra POMPER | The United Kingdom | Consumer Representative (Parent) |
| Amanda GWEE | Australia | Paediatrician, Infectious Diseases Physician and Clinical Pharmacologist |
| Amy LOVELL | New Zealand | Dietitian |
| Andrew GENNERY | The United Kingdom | Consultant Paediatrician |
| Annette HILL | The United Kingdom | Nurse Practitioner |
| Arjan LANKESTER | The Netherlands | Paediatric Immunologist/Infectiologist |
| Ben GELBART | Australia | Physician and Intensive Care Specialist |
| Eileen VAN DER STOEP | The Netherlands | Pharmacist |
| Elizabeth WILLIAMS | Australia | Pharmacist |
| Gabrielle HAEUSLER | Australia | Infectious Diseases Physician |
| Hilda MEKELENKAMP | The Netherlands | Specialist HSCT Nurse |
| Kanchan RAO | The United Kingdom | Consultant Paediatrician |
| Karen NICHOLSON | The United Kingdom | Nurse Practitioner |
| Katherine COLMAN | Australia | Paediatric Oncologist |
| Kavitha ASOKAN | The United Kingdom | Consumer Representative (Parent) |
| Lisa OTT DE BRUIN | The Netherlands | Paediatrician |
| Lori CHAIT-RUBINEK | Australia | Consumer Representative (Parent) |
| Lorna MCLEMAN | Australia | Paediatric Physician |
| Lucy TIMMINS | The United Kingdom | Consumer Representative (Parent) |
| Mary SLATTER | The United Kingdom | Consultant Paediatrician |
| Nader ELOSHAIKER | Australia | Consumer Representative (Parent) |
| Niaz CHOUDHURY | The United Kingdom | Consumer Representative (Parent) |
| Rachel CONYERS | Australia | Paediatric Oncologist  Project Principle Investigator |
| Rishya NADESWARAN | The United Kingdom | Consumer Representative (Parent) |
| Roxanne DYAS | Australia | Pharmacist |
| Shivanthan SHANTHIKUMAR | Australia | Paediatric Respiratory Physician |
| Suzannah KINSELLA | The United Kingdom | James Lind Alliance (JLA) Adviser |
| Theresa COLE | Australia | Paediatric Immunologist |
| Timothy PRESTIDGE | New Zealand | Consultant Physician and Pathologist |
| Vanessa CLIFFORD | Australia | Clinical Microbiologist |

**Supplementary 7:** **James Lind Alliance Priority Setting Partnership Steering Committee Members.** Abbreviations JLA: James Lind Alliance.
